# Hepatitis C Virus Infection and Risk of Non-Hodgkin’s Lymphoma: A Case-control Study in Mainland China

**DOI:** 10.1101/2023.09.14.23295415

**Authors:** Miao Hao, Wenjun Wang, Rui Lu, Yixin Liu, Fengping Wu, Yaping Li, Xiaoli Jia, Shuangsuo Dang, Xin Zhang

## Abstract

In order to evaluate the association between hepatitis C virus (HCV) and non-Hodgkin’s lymphoma (NHL) in Mainland China, we conducted a hospital case-control study. Cases were newly diagnosed patients with NHL between 2009 and 2015 admitted to the Second Affiliated Hospital of Xi’an Jiaotong University. Controls were gender and age matched patients admitted to the same hospital due to acute conditions. A total of 694 cases and controls (1 : 1 ratio) were identified. Anti-HCV prevalence was 1.7% in NHL cases and 2.3% in controls (6/347 *vs*. 8/347, OR = 0.75, 95% CI 0.26-2.17). Such prevalence in NHL cases (1.7%; 95% CI 0.6%-3.7%) does not differ from that in the Xi’an population (1.9%) or that in the general Chinese population (1.3%). HCV-RNA positivity was 0.9% in both NHL cases and controls using the available data (OR = 1.00, 95% CI 0.20-4.99). Subgroup analyses by histologic type of NHL or Epstein-Barr virus infection also showed no associations between anti-HCV and NHL. In conclusion, HCV is not associated with NHL risk in Mainland China, suggesting that the role of HCV in the occurrence of NHL is not significant in this population.

## Introduction

Hepatitis C virus (HCV) is an important public health threat, with an estimated 115 million people infected worldwide (1). In Mainland China, the prevalence of HCV infection was 1.3%, equating to approximately 15 million persons positive for antibodies to HCV (anti-HCV) (1). The causal effect of HCV infection on hepatitis, cirrhosis, and hepatocellular carcinoma is well defined. But the etiological associations of HCV and extrahepatic diseases are not that strong (2).

Non-Hodgkin’s lymphoma (NHL) is a heterogeneous malignant disease arising from lymphoid tissues. Like many other malignant diseases, environmental, genetic, and infectious factors contribute to the carcinogenesis of NHL. Among them, infectious agents such as Epstein-Barr virus (EBV), human T-cell leukemia virus, human immunodeficiency virus, human herpesvirus-8, and *Helicobacter pylori* are associated with certain subtypes of NHL (3). HCV, which infects hepatocytes and induces carcinogenesis of hepatocytes, also infects lymphocytes and replicates in them (4,5). This may induce error-prone DNA polymerases and produce high levels of reactive oxygen species in lymphocytes, thus enhancing mutation frequency and causing cellular DNA damage, respectively (6-8). In addition, the binding of HCV antigens and lymphocyte receptors activates proliferation signals (9-12). Actually, most published epidemiological studies found a significantly higher prevalence of HCV infection in patients with NHL than control populations; while other studies, especially conducted in areas with low rates of HCV infection, found weak or no association (13-20).

Currently, studies addressing the association of HCV infection with NHL risk are mainly conducted in Western people. Rare studies have addressed the issue in Chinese people. One study showed that HCV infection is associated with increased NHL risk in Taiwan (13), a area where HCV prevalence is higher than that in Mainland China (4.4% *vs*. 1.3%) (1). Therefore, we conducted a case-control study to further elucidate the issue in Mainland China.

## Material and methods

### Study population

This study is a retrospective case-control study. Cases with NHL were those consecutively admitted between July 2009 and February 2015 to the Second Affiliated Hospital of Xi’an Jiaotong University in Xi’an, China. Patients with a minimum age of 18 were eligible if they were newly diagnosed with NHL based on pathology. Patients with evidence of HIV infection, history of other malignancies or organ transplantation were excluded. Patients without testing for Anti-HCV infection were also excluded. Subtypes of NHL were classified according to the World Health Organization’s criteria (21).

Control subjects were randomly sampled from inpatient patients at the same hospital where cases were enrolled. They were frequency-matched to cases by gender and age (within ten years) in an 1 : 1 ratio. We selected patients admitted for acute conditions (e.g. trauma, acute appendicitis, etc) as controls since HCV positivity and these conditions are likely to be independent. We excluded those with chronic conditions (e.g. diabetes, cardiovascular and cerebrovascular diseases, malignant diseases, and autoimmune diseases, etc) because: (1) they might be associated with HCV infection (2); (2) patients with chronic diseases might change their lifestyle (22). Data were independently extracted from medical charts by two authors. Authors had no access to identifying information during or after data collection. As this was a retrospective study based on medical charts and all data were analyzed anonymously, informed consent was not obtained from participants. The study was approved by the ethics committee of the Second Affiliated Hospital of Xi’an Jiaotong University and was conducted according to the principles expressed in the Declaration of Helsinki.

### Laboratory assays

A specimen of whole blood was collected during hospitalization for each case and control and was centrifuged at 1500 g for 10 min to obtain serum. Samples were tested within 12 hr from collection or were stored at 4 °C and tested the next day.

Anti-HCV antibodies were tested using chemiluminescent microparticle immunoassay (ARCHITECT Anti-HCV; Abbott, Wiesbaden, Germany), a variation of the enzyme immunoassay. The assay detects antibodies to putative structural and nonstructural proteins of the HCV genome. Its sensitivity and specificity are estimated to be 99.60% and 99.10%, respectively. Anti-HCV positive samples were tested for serum HCV-RNA using HCV-RNA Quantitative Fluorescence Diagnostic Kit (Sansure Biotech, Changsha, China) with a lower limit of detection of 25 IU/mL.

### Statistical analysis

Characteristics of cases and controls were summarized as numbers and percentages and compared using Chi-square test. Odds ratio (OR) and corresponding 95% confidence interval (95% CI) were calculated to evaluate the risk of NHL for anti-HCV positivity. A sensitivity analysis was performed regarding HCV-RNA. Subgroup analyses of the association between anti-HCV and NHL were performed for histological subtype and EBV infection. The prevalence of anti-HCV positivity was compared to literature results using the exact binomial method (1,23). The statistical analyses were performed using STATA software (version 12.0, StataCorp LP, College Station, Texas, USA).

## Results

A total of 352 NHL cases were identified. Excluding 5 younger than 18 years old, 347 were included in the analysis. After excluding those without testing for anti-HCV or younger than 18 years old and matching for gender and age, 347 from 618 controls with acute conditions in the same hospital were included. Of these, 59.7% were admitted for abdominal surgery, 13.0% for thoracic surgery, 16.1% for brain surgery, 5.5% for urinary surgery, and 5.8% for eye surgery. The distribution of NHL cases and controls by gender, age, race, and residence was shown in Table 1. Race and residence were not significantly associated with NHL risk.

**Table 1.** Characteristics of study participants.

|  | Cases n=347 (%) | Controls n=347 (%) | *P* value<sup>1)</sup> | OR (95% CI) |
| --- | --- | --- | --- | --- |
| Gender |  |  |  |  |
| Male | 230 (66.3%) | 230 (66.3%) |  |  |
| Female | 117 (33.7%) | 117 (33.7%) |  |  |
| Age | 56 (18-87)<sup>2)</sup> | 55 (18-85)<sup>2)</sup> |  |  |
| 18-20 | 5 (1.4%) | 5 (1.4%) |  |  |
| 20-30 | 26 (7.5%) | 26 (7.5%) |  |  |
| 30-40 | 32 (9.2%) | 32 (9.2%) |  |  |
| 40-50 | 58 (16.7%) | 58 (16.7%) |  |  |
| 50-60 | 92 (26.5%) | 92 (26.5%) |  |  |
| 60-70 | 87 (25.1%) | 87 (25.1%) |  |  |
| 70-80 | 42 (12.1%) | 42 (12.1%) |  |  |
| >= 80 | 5 (1.4%) | 5 (1.4%) |  |  |
| Race |  |  |  |  |
| Han | 341 (98.3%) | 342 (98.6%) |  | 1.00<sup>3)</sup> |
| Hui | 6 (1.7%) | 5 (1.4%) | 0.76 | 1.20 (0.36-3.98) |
| Residence |  |  |  |  |
| City | 188 (54.2%) | 165 (47.6%)<sup>4)</sup> |  | 1.00<sup>3)</sup> |
| Rural | 159 (45.8%) | 180 (51.9%)<sup>4)</sup> | 0.10 | 0.78 (0.58-1.05) |

CI: confidence interval; OR: odds ratio.
<sup>1)</sup>: P value from Chi-square test.
<sup>2)</sup>: Presented as median (range).
<sup>3)</sup>: Reference category.
<sup>4)</sup>: Residence information is not available in two controls.

Anti-HCV prevalence was 1.7% in NHL cases (6/347) and 2.3% in controls (8/347) (OR = 0.75, 95% CI 0.26-2.17). Such prevalence in NHL cases (1.7%; 95% CI 0.6%-3.7%) does not differ from that in the Xi’an population (1.9%) or that in the general Chinese population (1.3%) (1,23). Subgroup analyses by histologic type of NHL or EBV infection also showed no associations between anti-HCV and NHL risk (Table 2).

**Table 2.** Association between HCV infection and NHL risk.

| Category | Anti-HCV positive (%) | Anti-HCV negative (%) | OR (95% CI) |
| --- | --- | --- | --- |
| **Controls (n=347)** | 8 (2.3%) | 339 (97.7%) | 1.00<sup>1)</sup> |
| **NHL cases (n=347)** | 6 (1.7%) | 341 (98.3%) | 0.75 (0.26-2.17) |
| **Cases by histologic type** |  |  |  |
| **All B-cell (n=190)** | 3 (1.6%) | 187 (98.4%) | 0.68 (0.18-2.59) |
| **Diffuse large cell (n=76)** | 1 (1.3%) | 75 (98.7%) | 0.57 (0.07-4.59) |
| **Follicular (n=12)** | 0 (0%) | 12 (100%) | - |
| **Other types (n=102)** | 2 (2.0%) | 100 (98.0%) | 0.85 (0.18-4.06) |
| **All T-cell (n=114)** | 3 (2.6%) | 111 (97.4%) | 1.15 (0.30-4.39) |
| **Unknown (n=43)** | 0 (0%) | 43 (100%) | - |
| **Cases by** |  |  |  |
| **EBV infection (n=63)** | 2 (3.2%) | 61 (96.8%) | 1.39 (0.29-6.70) |
| **No EBV infection (n=221)** | 3 (1.4%) | 218 (98.6%) | 0.58 (0.15-2.22) |
| **Unknown (n=63)** | 1 (1.6%) | 62 (98.4%) | 0.68 (0.08-5.56) |

CI: confidence interval; EBV: Epstein-Barr virus; HCV: hepatitis C virus; NHL: non-Hodgkin's lymphoma; OR: odds ratio.
<sup>1)</sup>: Reference category.

All the 6 NHL cases positive for anti-HCV were tested for serum HCV-RNA and 3 were positive. Of the 8 controls positive for anti-HCV, 5 were tested for serum HCV-RNA and 3 were positive. Thus, HCV-RNA positivity was 0.9% in both NHL cases (3/347) and controls (3/347) using the available data (OR = 1.00, 95% CI 0.20-4.99), or 0.9% in NHL cases (3/347) and 1.7% in controls (6/347) assuming that the 3 controls positive for anti-HCV but without HCV-RNA detection were positive for HCV-RNA (OR = 0.50, 95% CI 0.12-2.00). Such viremia prevalence in NHL cases (0.9%; 95% CI 0.2%-2.5%) does not differ from that in the Xi’an population (1.9%) or that in the general Chinese population (0.8%).

## Discussion

Serum anti-HCV antibodies are indicative of past or active HCV infection and serum HCV-RNA is the indicator of persistent infection. In this study, we found an 1.7% prevalence of past or active, and 0.9% prevalence of active HCV infection in patients with NHL. Such prevalences do not differ from those in controls, in the Xi’an population, or in the general Chinese population (1,23).

Over the last two decades, large number of epidemiologic studies have investigated the association between HCV infection and NHL risk in the Western population (15-20). Although most have found positive associations, the strength of the association greatly varies between and within countries. In countries with high background prevalence of HCV (e.g., Italy and Egypt), the ORs are as high as 2-3 on average and even more than 10 in some studies, while in low prevalence countries (e.g., Canada, Scandinavia, and UK) the association is weak or not evident (15-20). Recently, a cohort study performed by Su *et al* in Taiwan, an area with high HCV prevalence (4.4%), showed 2-fold NHL risk in Chinese population (13). As for the Chinese population in a low prevalence area, our study found no significant association. Together, the association varies with HCV prevalence in the Chinese population just as that in the Western population.

The above phenomenon can also be seen in the association of HBV infection and NHL risk (24). The association strength decreases as the exposure intensity decreases. Confounding factors are prone to hide the real association if the association is not strong enough and the exposure intensity is low. By enlarging sample size, statistically significant ORs may be achieved in areas with low HCV prevalence. However, there are often mild. The sample size in our study is not comparable with recently published studies involving large areas (13,14,17). However, it is large enough compared with early studies. At least, the findings from our study suggest that the role of HCV in the development of NHL is not significant in Mainland China.

Previous studies indicate that HCV prevalence is associated with B-cell NHL. Among B-cell NHL, marginal zone lymphoma, diffuse large B-cell lymphoma, and lymphoplasmacytic lymphoma were identified as most closely related to HCV in previous studies (16-20). However, in our study, the case number in each subgroup was too small to draw a conclusion. We also failed to detect an association of HCV infection with EBV-positive NHL or EBV-negative NHL. Large samples are needed in future studies when evaluating each subtype in the Chinese population.

Some molecular mechanisms have been proposed to support HCV-associated lymphomagenesis. These involve continuous external stimulation of lymphocyte receptors by viral antigens, consecutive proliferation and oncogenic effects mediated by intracellular viral proteins, and permanent genetic lymphocyte damage (16). Nevertheless, these mechanisms are still poorly understood and need further confirmation. More robust evidence is needed to elucidate whether HCV causes NHL. Besides humans, natural infection of HCV is limited to chimpanzees and it may take a long time to develop lymphoma in them. Therefore, it would be difficult to prove the etiology role of HCV in animal models. Controlled intervention studies to observe whether eradication of HCV can decrease NHL risk are feasible and will supply more solid evidence.

The strength of the current study lies in the consistent results from analyses by indicator of HCV infection (anti-HCV and HCV-DNA) and from analyses using reference rates of HCV prevalence from three different populations. Missing data of HCV-DNA in controls positive for anti-HCV did not affect the results, either. However, several limitations are worth mentioning. Firstly, like most retrospective studies addressing the association of HCV infection with NHL risk, due to the nature of study design, risk factors such as immunodeficiency, human T-cell leukemia virus, human herpesvirus-8, and *Helicobacter pylori* were not available in our study. They may affect the results especially when the interested associations are not strong and the HCV prevalence is low. Secondly, risk factors of HCV infection and HCV genotype were not available, either, though they are unrelated to NHL risk according to previous studies (17-20). Thirdly, only participants positive for anti-HCV were considered for HCV-RNA detection. However, this is not likely to change the results because of the high specificity and sensitivity of anti-HCV assay. What’s more, lymphomagenesis is a long-term process so that patients during the window phase of HCV infection (a period of usually several months, during which HCV replication can be detected and anti-HCV IgG antibodies have not developed) have little effect on the association results.

In conclusion, our study shows that HCV infection is not associated with NHL risk in Mainland China, a low HCV prevalence area. This finding suggests that the role of HCV infection in the occurrence of NHL is not significant in this population.

## Data Availability

Data supporting the findings of this study are not publicly available due to privacy or ethical restrictions and are available upon request to the corresponding author.

## Conflict of interest

None to declare.

## Notes

### Competing Interest Statement

The authors have declared no competing interest.

### Funding Statement

This study did not receive any funding

### Author Declarations

The study was approved by the ethics committee of the Second Affiliated Hospital of Xi'an Jiaotong University

## Reference

1. Gower E, Estes C, Blach S, et al. Global epidemiology and genotype distribution of the hepatitis C virus infection. J Hepatol. 2014; 61:S45–57.

2. Jacobson IM, Cacoub P, Dal Maso L, et al. Manifestations of chronic hepatitis C virus infection beyond the liver. Clin Gastroenterol Hepatol. 2010; 8:1017–29.

3. Fisher SG, Fisher RI. The epidemiology of non-Hodgkin’s lymphoma. Oncogene. 2004; 23:6524–34.

4. Bare P, Massud I, Parodi C, et al. Continuous release of hepatitis C virus (HCV) by peripheral blood mononuclear cells and B-lymphoblastoid cell-line cultures derived from HCV-infected patients. J Gen Virol. 2005; 86:1717–27.

5. Pal S, Sullivan DG, Kim S, et al. Productive replication of hepatitis C virus in perihepatic lymph nodes in vivo: implications of HCV lymphotropism. Gastroenterology. 2006; 130:1107–16.

6. Machida K, Cheng KT, Lai CK, et al. Hepatitis C virus triggers mitochondrial permeability transition with production of reactive oxygen species, leading to DNA damage and STAT3 activation. J Virol. 2006; 80:7199–207.

7. Machida K, Cheng KT, Sung VM, et al. Hepatitis C virus infection activates the immunologic (type II) isoform of nitric oxide synthase and thereby enhances DNA damage and mutations of cellular genes. J Virol. 2004; 78:8835–3.

8. Machida K, Cheng KT, Sung VM, et al. Hepatitis C virus induces a mutator phenotype: enhanced mutations of immunoglobulin and protooncogenes. Proc Natl Acad Sci U S A. 2004; 101:4262–7.

9. Dai B, Chen AY, Corkum CP, et al. Hepatitis C virus upregulates B-cell receptor signaling: a novel mechanism for HCV-associated B-cell lymphoproliferative disorders. Oncogene. 2015; 35:2979.

10. Kasama Y, Mizukami T, Kusunoki H, et al. B-cell-intrinsic hepatitis C virus expression leads to B-cell-lymphomagenesis and induction of NF-kappaB signalling. PLoS One. 2014; 9:e91373.

11. Feldmann G, Nischalke HD, Nattermann J, et al. Induction of interleukin-6 by hepatitis C virus core protein in hepatitis C-associated mixed cryoglobulinemia and B-cell non-Hodgkin’s lymphoma. Clin Cancer Res. 2006; 12:4491–8.

12. Rosa D, Saletti G, De Gregorio E, et al. Activation of naive B lymphocytes via CD81, a pathogenetic mechanism for hepatitis C virus-associated B lymphocyte disorders. Proc Natl Acad Sci U S A. 2005; 102:18544–9.

13. Su TH, Liu CJ, Tseng TC, et al. Hepatitis C viral infection increases the risk of lymphoid-neoplasms: A population-based cohort study. Hepatology. 2016; 63:721–30.

14. Abe SK, Inoue M, Sawada N, et al. Hepatitis B and C virus infection and risk of lymphoid malignancies: A population-based cohort study (JPHC Study). Cancer Epidemiol. 2015; 39:562–6.

15. Aledavood SA, Ghavam-Nasiri MR, Ghaffarzadegan K, et al. Hepatitis-C Infection Incidence Among the non-Hodgkin’s B-cell Lymphoma Patients in the Northeast of Iran. Iran J Cancer Prev. 2014; 7:147–51.

16. Peveling-Oberhag J, Arcaini L, Hansmann ML, et al. Hepatitis C-associated B-cell non-Hodgkin lymphomas. Epidemiology, molecular signature and clinical management. J Hepatol. 2013; 59:169–77.

17. de Sanjose S, Benavente Y, Vajdic CM, et al. Hepatitis C and non-Hodgkin lymphoma among 4784 cases and 6269 controls from the International Lymphoma Epidemiology Consortium. Clin Gastroenterol Hepatol. 2008; 6:451–8.

18. Dal Maso L, Franceschi S. Hepatitis C virus and risk of lymphoma and other lymphoid neoplasms: a meta-analysis of epidemiologic studies. Cancer Epidemiol Biomarkers Prev. 2006; 15:2078–85.

19. Matsuo K, Kusano A, Sugumar A, et al. Effect of hepatitis C virus infection on the risk of non-Hodgkin’s lymphoma: a meta-analysis of epidemiological studies. Cancer Sci. 2004; 95:745–52.

20. Gisbert JP, Garcia-Buey L, Pajares JM, et al. Prevalence of hepatitis C virus infection in B-cell non-Hodgkin’s lymphoma: systematic review and meta-analysis. Gastroenterology. 2003; 125:1723–32.

21. Jaffe ES, Harris NL, Stein H, et al. Pathology and genetics of tumours of haematopoietic and lymphoid tissues. IARC Press; 2001.

22. Talamini R, Montella M, Crovatto M, et al. Non-Hodgkin’s lymphoma and hepatitis C virus: a case-control study from northern and southern Italy. Int J Cancer. 2004; 110:380–5.

23. Qu M, An Z, Li L. Epidemiological status of HBV and HCV infection in Xi’an area. Ji Bing Jian Ce Yu Kong Zhi. 2012; 6:73–4. Chinese.

24. Qi Z, Wang H, Gao G. Association of risk of non-Hodgkin’s lymphoma with hepatitis B virus infection: a meta-analysis. Int J Clin Exp Med. 2015; 8:22167–74.

